# Characterizing all-cause excess mortality patterns during COVID-19 pandemic in Mexico

**DOI:** 10.1101/2021.03.02.21252763

**Authors:** Sushma Dahal, Juan M. Banda, Ana I. Bento, Kenji Mizumoto, Gerardo Chowell

## Abstract

**Background:** Low testing rates and delays in reporting hinder the estimation of the mortality burden associated with the COVID-19 pandemic. During a public health emergency, estimating all cause excess deaths above an expected level of death can provide a more reliable picture of the mortality burden. Here, we aim to estimate the absolute and relative mortality impact of COVID-19 pandemic in Mexico.

**Methods:** We obtained weekly mortality time series due to all causes for Mexico, and by gender, and geographic region from 2015 to 2020. We also compiled surveillance data on COVID-19 cases and deaths to assess the timing and intensity of the pandemic and assembled weekly series of the proportion of tweets about ‘death’ from Mexico to assess the correlation between people’s media interaction about ‘death’ and the rise in pandemic deaths. We estimated all-cause excess mortality rates and mortality rate ratio increase over baseline by fitting Serfling regression models and forecasted the total excess deaths for Mexico for the first four weeks of 2021 using the generalized logistic growth model.

**Results:** We estimated the all-cause excess mortality rate associated with the COVID-19 pandemic in Mexico in 2020 at 26.10 per 10,000 population, which corresponds to 333,538 excess deaths. Males had about 2-fold higher excess mortality rate (33.99) compared to females (18.53). Mexico City reported the highest excess death rate (63.54) and RR (2.09) compared to rest of the country (excess rate=23.25, RR=1.62). While COVID-19 deaths accounted for only 38.64% of total excess deaths in Mexico, our forecast estimate that Mexico has accumulated a total of ∼61610 [95% PI: 60003, 63216] excess deaths in the first four weeks of 2021. Proportion of tweets was significantly correlated with the excess mortality (ρ=0.508 [95% CI: 0.245, 0.701], p-value=0.0004).

**Conclusion:** The COVID-19 pandemic has heavily affected Mexico. The lab-confirmed COVID-19 deaths accounted for only 38.64% of total all cause excess deaths (333,538) in Mexico in 2020. This reflects either the effect of low testing rates in Mexico, or the surge in number of deaths due to other causes during the pandemic. A model-based forecast indicates that an average of 61,610 excess deaths have occurred in January 2021.

## Background

SARS-CoV-2 continues to spread unabated in many parts of the world. In particular, Latin American countries are being heavily affected by the COVID-19 pandemic with a total of 22,467,574 cases including 709,062 deaths as of March 12, 2021 [1]. Mexico, one of the highly populated countries in the world with approximately 42% of the people living in poverty [2], documented the first imported COVID-19 case on 27^th^ February, 2020 and currently ranks 3rd in the world in terms of numbers of COVID-19 reported deaths, with a total of 192,488 recorded deaths (7.33% of total deaths globally) as of March 12, 2021 [1]. Delays in the implementation of social distancing interventions, mixed reactions towards the stay-at-home order recommendations, and phased reopening of the country have facilitated sustained transmission of COVID-19 in Mexico [3].

Mexico has one of the lowest per-capita COVID-19 testing rates in the world with about 17 tests per 1000 people in total [4]. The low testing rates, compounded by reporting delays, hinders the estimation of the mortality burden associated with the COVID-19 pandemic based on surveillance data alone. Instead, a more reliable picture of the effect of COVID-19 pandemic on mortality can be derived by estimating excess deaths above a baseline or expected level of death [5, 6]. These estimates can provide information about the deaths that are directly or indirectly attributed to the pandemic [6]. Indeed, some deaths could be misclassified as COVID-19 deaths, or some could be occurring in the context of overburdened health care systems. Thus, tracking all-cause mortality in near real time can help assess whether excess deaths are occurring during a specific period of time and spatial area [6].

Here we report our estimates of the absolute and relative mortality impact of the COVID-19 pandemic in Mexico using cyclical Serfling regression models together with publicly available weekly all-cause mortality data from 2015 to 2020 by gender and for Mexico City and other areas of Mexico. Further, we collected and analyzed weekly twitter data from Mexico about ‘deaths’ during the COVID-19 pandemic in correlation with the excess all-cause death rate and COVID-19 death rate. We supplemented our analyses with social media data from Twitter, which has been found useful to interpret epidemiological trends [7]. In prior work, Google Trends, Wikipedia searches, and Twitter data have been used for predicting COVID-19 deaths [8]. Taking this data fusion approach even further, other researchers have used Twitter data, alongside smart thermometer data, up-to-date clinician search logs as well as Apple and Cuebiq mobility indices to build near-real time early warning systems for COVID-19 [9].

We also generated predictions of excess mortality for the first 4 weeks of January 2021 using generalized logistic growth model [10].

## Methods

*Data:* We obtained weekly all-cause death counts based on epidemiological weeks for Mexico which were also stratified by gender and geographic region from January to December 2020 as well as for the preceding 5 years (2015-2019) in order to establish a baseline mortality level [11]. We accessed publicly available weekly mortality data available from National Institute of Statistics and Geography (INEGI) for the years from 2015 to 2018, and data available from National Population Registry (RENAPO) for the years 2019 and 2020 [11]. The last week of December 2020, also includes first 2 days of January 2021. To gauge the timing and relative intensity of the pandemic in Mexico, we examined surveillance data characterizing the weekly number of laboratory-confirmed COVID-19 cases and deaths, which were obtained from the official website of the Mexican Ministry of Health through the Directorate General of Epidemiology [12]. Population size estimates used to calculate mortality rates were obtained from National Population Council (CONAPO) of Mexico [13].

### Statistical analysis

To investigate and quantify the mortality pattern associated with the COVID-19 pandemic in Mexico, we estimated excess all-cause mortality rates per 10,000 population at the national level and for Mexico City, and other areas of Mexico and by gender. The excess death rate corresponds to the overall mortality rate above a seasonal baseline of the expected mortality rates in the absence of the COVID-19 pandemic using standard statistical methods [5, 14-17].

### Definition of pandemic periods and excess mortality estimation

We estimated the baseline mortality level by fitting cyclical Serfling regression models to all-cause deaths in non-COVID-19 period, after excluding data from March to December 2020. We included a combination of linear terms with sine and cosine terms describing time trend and seasonal change respectively, which is described by the following equation [5, 15]:

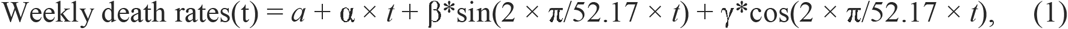

Where, *a* is the intercept and *t* represents the epidemic week.

Once a weekly baseline and 95% CI were established, periods of COVID-19 pandemic were defined as the weeks in 2020 where the observed all-cause mortality rate exceeded the upper 95% confidence limit of the baseline mortality level. The same pandemic period was used for estimating the total excess mortality rate for entire Mexico, Mexico City, Mexico excluding Mexico City, and gender specific excess mortality rates using established methodology [5, 14-17]. Excess all-cause mortality rate was defined as the difference between the observed and model adjusted baseline mortality rates for each week constituting the pandemic period. Negative excess mortality estimates were replaced by zeros in our analyses. Overall pandemic excess mortality attributed to all cause for total population, each gender group, Mexico City, and Mexico excluding Mexico City was calculated by summing the excess death rates across the pandemic weeks in 2020 [14, 16]. We also calculated the rate ratio (RR), the ratio of observed all-cause mortality rate during pandemic period to the model predicted baseline mortality level in the absence of COVID-19 for the given group.

### Twitter data analysis

We used a clean version of the publicly available dataset of tweets version 42 [18], the clean version of this dataset removes all re-tweets, keeping only directly initiated posts by users. We filtered all tweets, by removing all other languages via their ISO 639-1 language code, to only keep the tweets in Spanish (es) and those that originated from Mexico via its country code MX. Additionally, we removed tweets from news agencies and bot accounts. We used the following terms to subset the tweets per day: “muerto, muerta, fallecio, murio, deceso, fallecimiento, defunción, óbito, expiración, defuncion, obito, expiracion, perdio la vida, sin vida”. In English, these terms reflect the meanings “dead, deceased, died, death, expiration, lost life, lifeless”. We collected a total of 1,223,096 and 32,423,282 unique tweets reflecting death and total tweets respectively from March to December 2020. Next, we overlayed the curve of proportion of tweets on death out of total tweets in a given week over the mortality rate curve to inspect the relationship between the mortality rate and the proportion of tweets. We also calculated correlation coefficients between proportion of weekly tweets and the weekly excess death rate and the weekly COVID-19 death rate.

### Short term forecast of excess deaths

We used generalized logistic growth model (GLM) to predict excess deaths during the first four weeks of 2021. GLM characterizes epidemic growth by estimating (i) the intrinsic growth rate, *r* (ii) a dimensionless “deceleration of growth” parameter, *p* and (iii) the final epidemic size, *k*_0_. The deceleration parameter modulates the epidemic growth patterns including the sub-exponential growth (0< *p* <1), constant incidence (*p* =0) and exponential growth dynamics (*p* =1). The GLM model is given by the following differential equation:

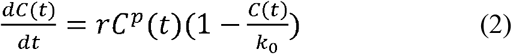

Where,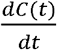 describes the incidence (of excess deaths) over time *t* and the cumulative number of excess deaths at time *t* is given by *C(t)*. To forecast the number of excess deaths during the first 4 weeks of 2021, we calibrated the model using weekly excess deaths during the last six weeks of 2020 (week starting from November 22 to the week starting from December 27, 2020). We utilized parametric bootstrapping approach with a Poisson error structure [10].

## Results

Between March 1, 2020 and January 2, 2021, as of surveillance data updated on February 26, 2021, a total of 1,364,557 laboratory-confirmed COVID-19 cases, and 128,886 COVID-19-related deaths were captured by the epidemiological surveillance system in Mexico. The national daily series of new cases and deaths due to COVID-19 are shown in Figure 1. The number of cases rapidly rises from April to July followed by a downward trend which again takes off from mid-September. A similar temporal pattern can also be gleaned from the time series of COVID-19 related deaths.

**Figure 1.**
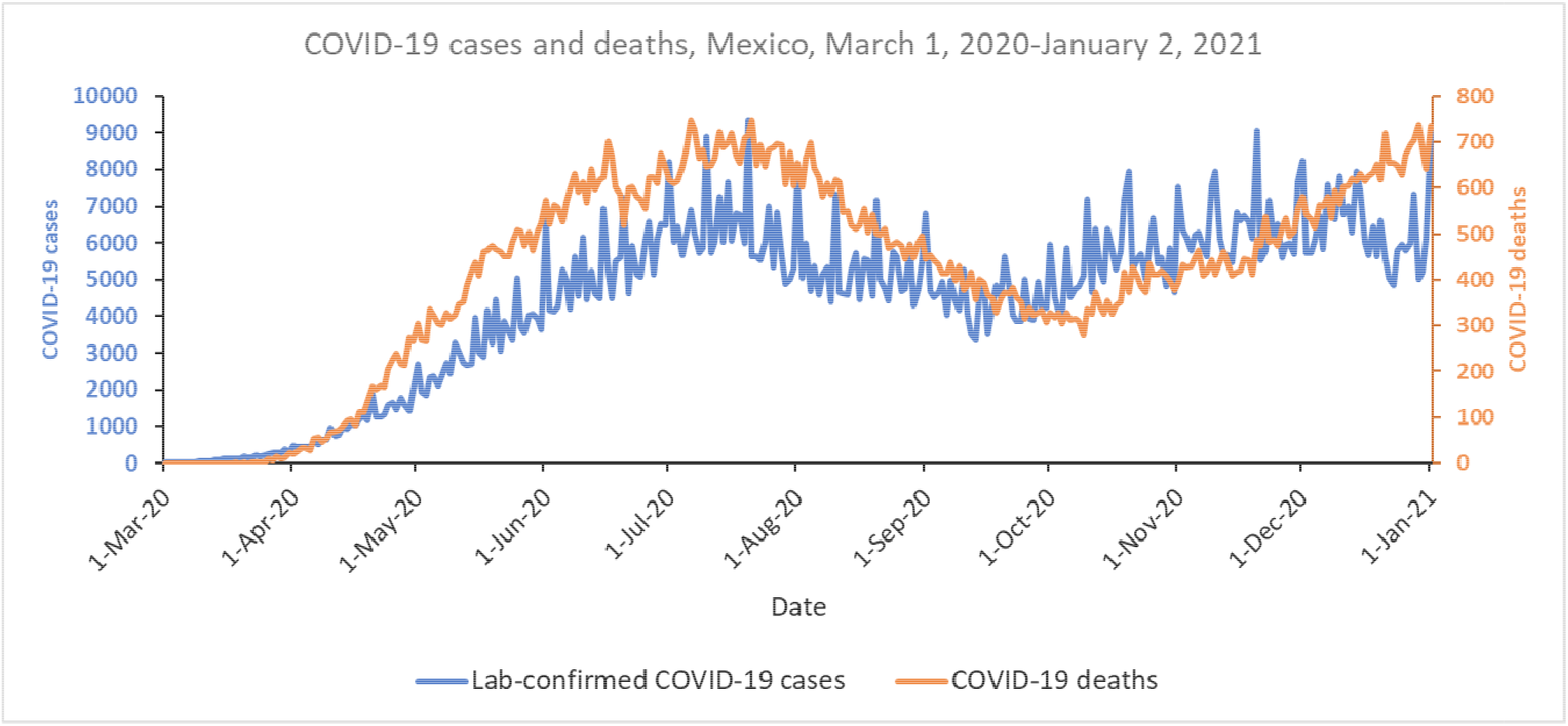
Daily series of new laboratory-confirmed COVID-19 cases and deaths in Mexico, from March 1, 2020-January 2, 2021

Out of total 44 weeks from March 1, 2020 to January 2, 2021, 38 weeks starting from week 16 (April 12-18, 2020) had the excess death rate greater than 0. The excess death rate peaked on week 29 (July 12-18, 2020) with the excess death rate of 1.01 per 10,000 population, and on week 53 (December 27, 2020-January 2, 2021) with the excess death rate of 1.06 per 10,000 population. The weekly timeseries of all-cause mortality rate per 10,000 population in Mexico is shown in (Figure 2). We found that peaks in all-cause death rates aligned with the peaks in COVID-19 laboratory-confirmed death rates captured by the surveillance system. The curve of weekly proportion of tweets from Mexico about death is overlaid with the mortality rate curve in Figure 2.

**Figure 2.**
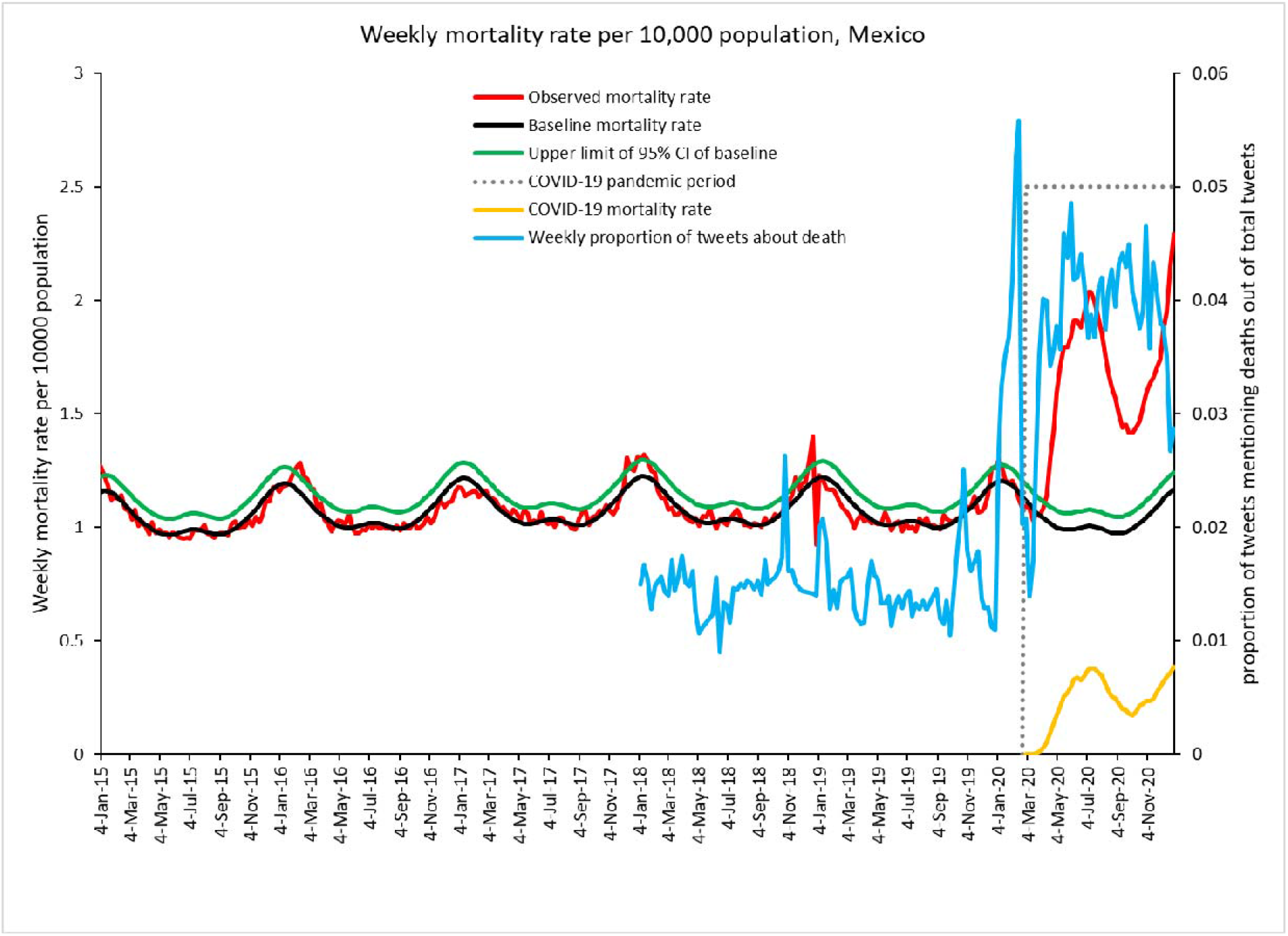
Mortality rate per 10,000 population, Mexico, 2015-2020. The red line is the weekly all-cause death rate. COVID-19 death rate curve is shown in yellow. Dotted lines highlight 2020 COVID-19 pandemic period. The Serfling seasonal regression model baseline (*black curve*) and corresponding upper limit of the 95% confidence interval of the baseline (*green curve*) are also shown. The weekly frequency of tweets about death is shown by blue curve. Excess all-cause mortality rate is the difference between the observed and model adjusted baseline mortality rates for each week where observed total all-cause mortality rate exceeded the upper 95% confidence limit of the baseline.

Twitter trends show engagement of people in Mexico with the hashtag terms (Figure 2). The trend of tweets with the hashtag term related to deaths (explained in the methods section) for the baseline years of 2018 and 2019 shows a sharp increase in the twitter chatter about deaths during the pandemic compared to the baseline years. There was a relatively weak but statistically significant correlation of the weekly proportion of tweets with the weekly excess mortality rate ρ=0.508 [95% CI: 0.245, 0.701], p-value=0.0004, and the weekly COVID-19 mortality rate ρ=0.526 [95% CI: 0.268, 0.714], p-value=0.0002, in the study period

The weekly timeseries of all-cause mortality rate per 10,000 population in the country of Mexico by gender are displayed in Figure 3.

**Figure 3.**
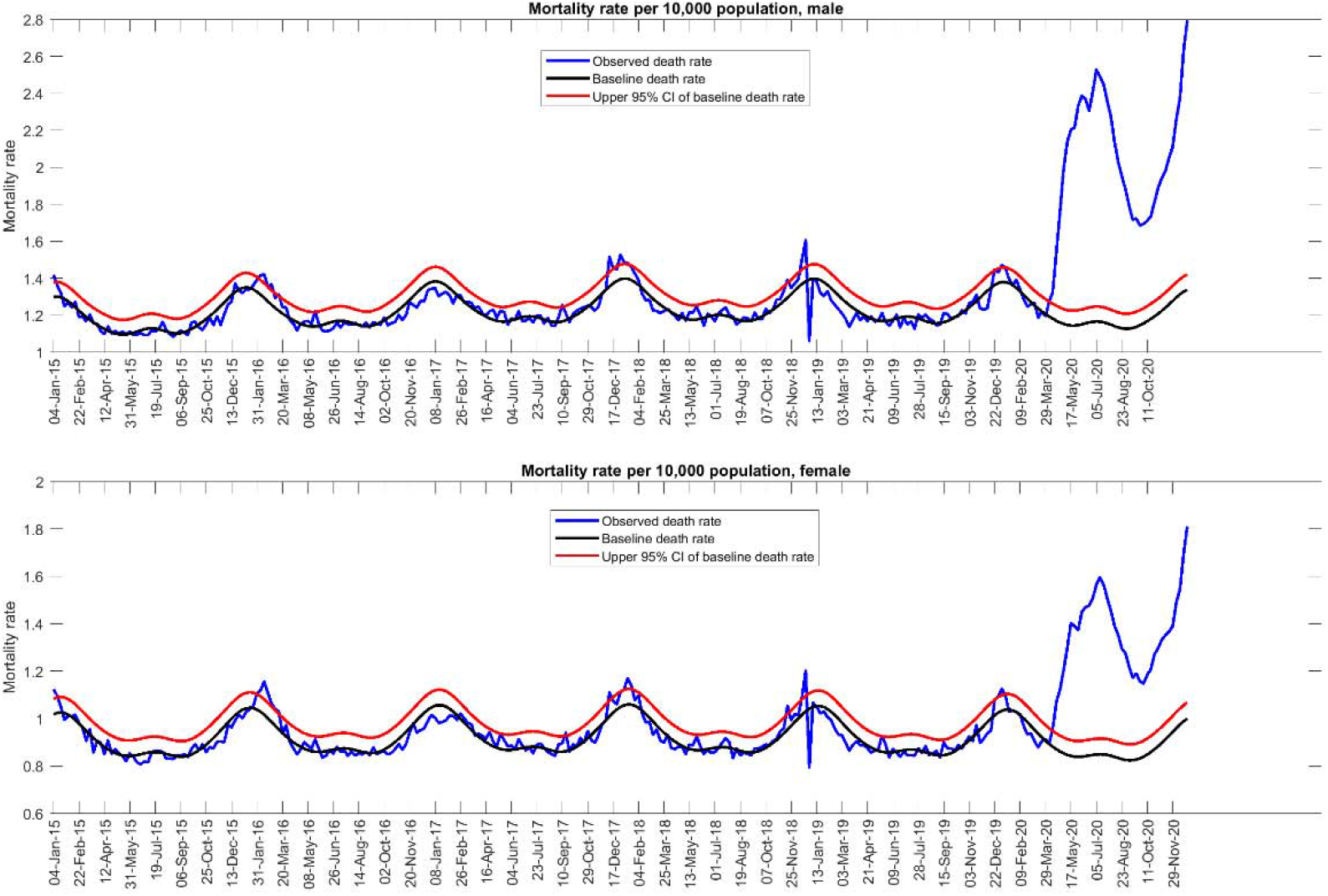
Mortality rate per 10,000 by gender, Mexico. Excess all-cause mortality rate is the difference between the observed and model adjusted baseline mortality rates for each week where observed total all-cause mortality rate exceeded the upper 95% confidence limit of the baseline in the country.

Similarly, the weekly timeseries of all-cause mortality rate per 10,000 population for Mexico City and for the rest of Mexico are shown in Figure 4.

**Figure 4.**
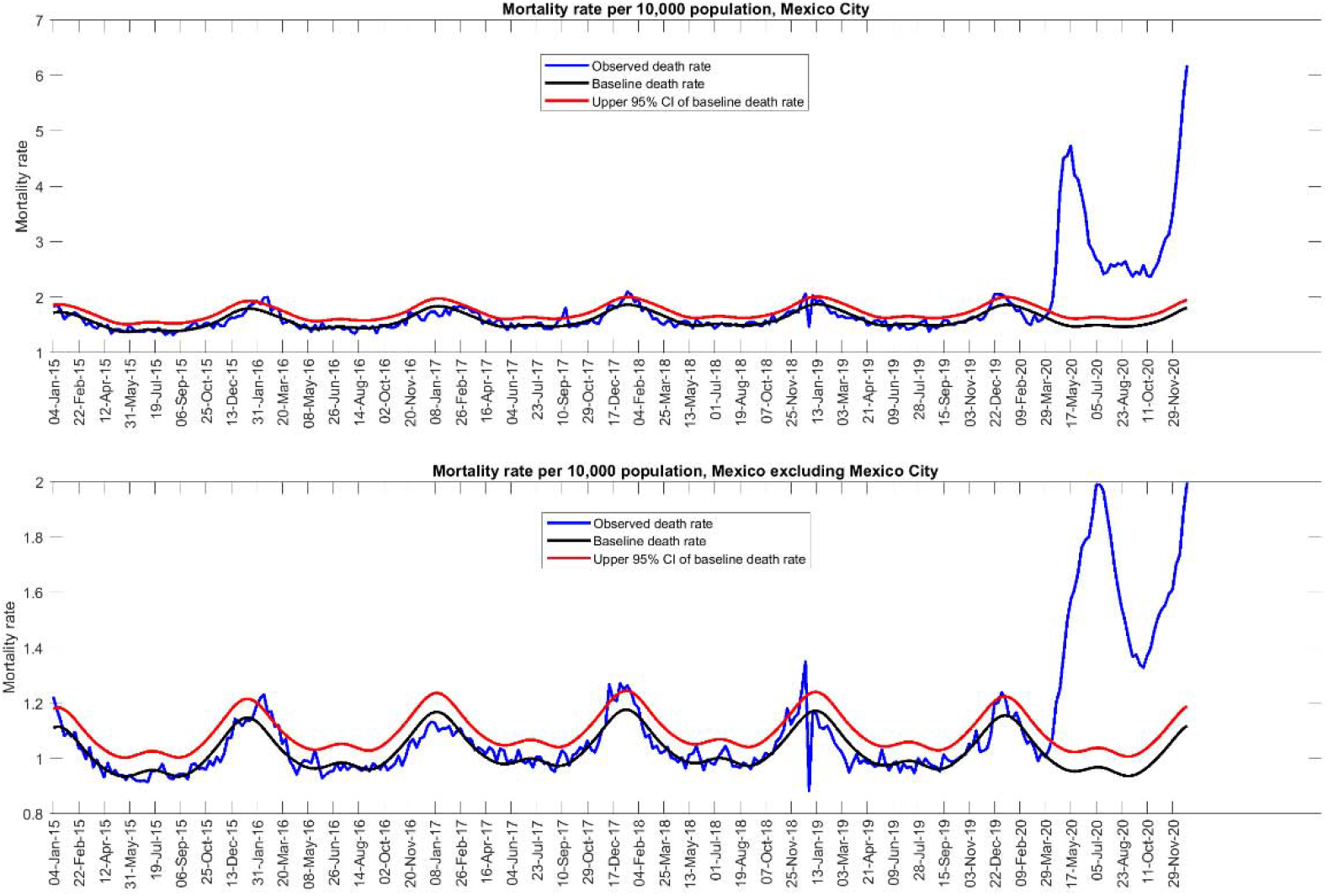
Mortality rate for Mexico City and Mexico excluding Mexico City. Excess all-cause mortality rate is the difference between the observed and model adjusted baseline mortality rates for each week where observed total all-cause mortality rate exceeded the upper 95% confidence limit of the baseline in the country.

In Table 1, we present the estimates of all-cause excess mortality rate per 10,000 population and the rate ratio estimates for each studied group, including the estimates at the national level. We estimated an excess death rate at 26.10 per 10,000 population in Mexico from March 1 to January 2, 2021. This corresponds to 333,538 excess deaths during the pandemic period. In the same period, a total of 128,886 lab-confirmed COVID-19 deaths corresponds to 38.64% of the total estimated excess deaths.

**Table 1.**
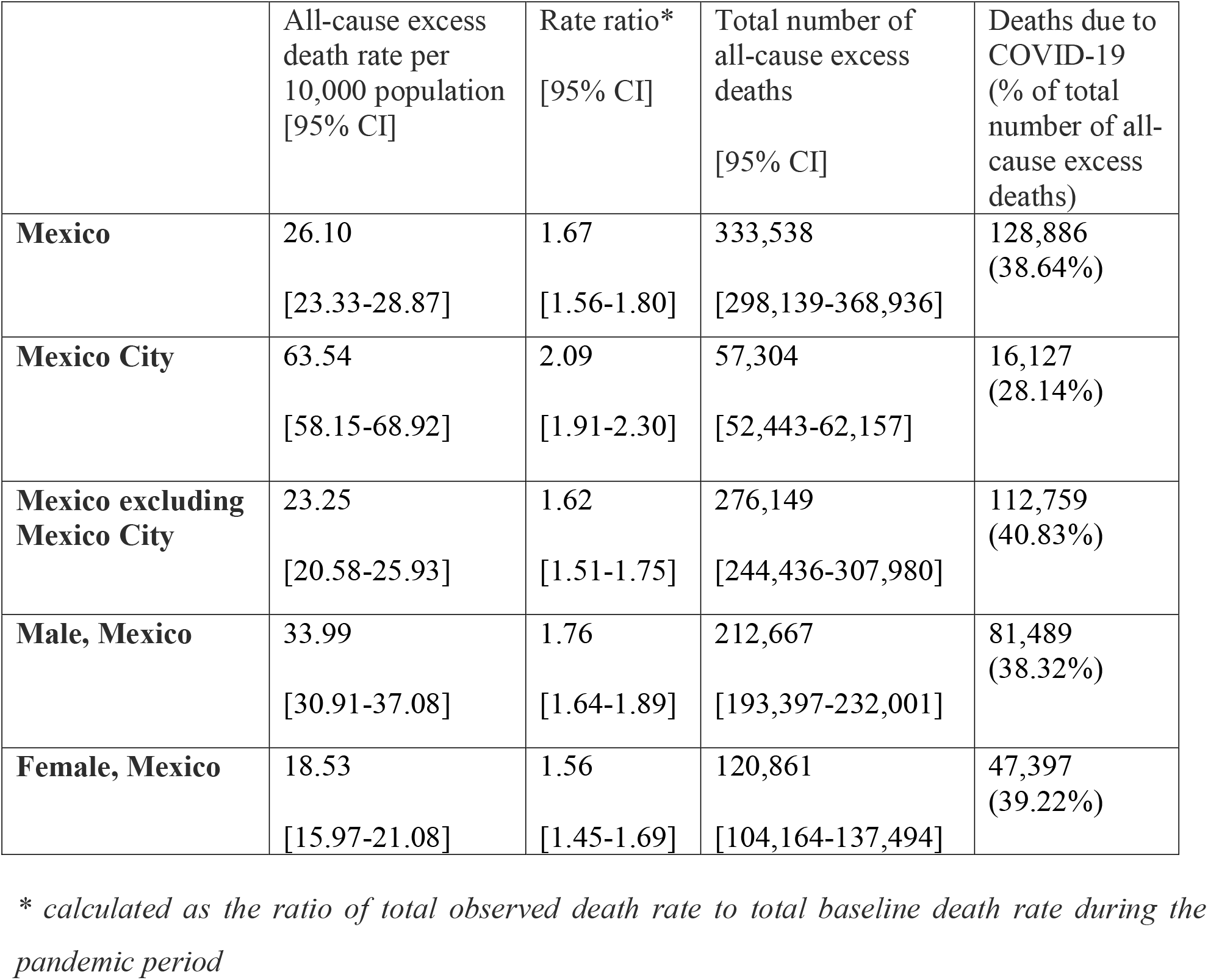
Estimates and their uncertainty for all-cause excess mortality rates per 10,000 population and RR during COVID-19 pandemic, Mexico, March 1, 2020- January 2, 2021.

The excess mortality rate in Mexico City (63.54) was about 2.7-fold higher than the rest of the Mexico (23.25) (proportion test, p-value <0.0001). Interestingly, COVID-19 deaths in Mexico City accounted for only 28.14% % of the total estimated excess deaths in Mexico City, compared to 40.83% in the rest of Mexico. Excess mortality rate among males nearly doubled the rate among females (proportion test, p-value <0.0001, and the proportion of COVID-19 deaths out of total excess deaths was similar, 38.32% among males and 39.22% among females (proportion test, p-value <0.0001).

Our estimates of both the absolute and relative excess mortality rate as measured by the rate ratio of observed vs baseline mortality rate was highest for Mexico City (2.09) compared to other areas of Mexico (1.62) and for males (1.76). The rate ratio (RR) at the national level was estimated at 1.67.

Finding from the average weekly forecast of excess deaths generated from the GLM model calibrated to weeks starting from November 22 to December 27, 2020 shows that Mexico had a total of ∼61610 excess deaths in first four weeks of 2021 (Figure 5).

**Figure 5.**
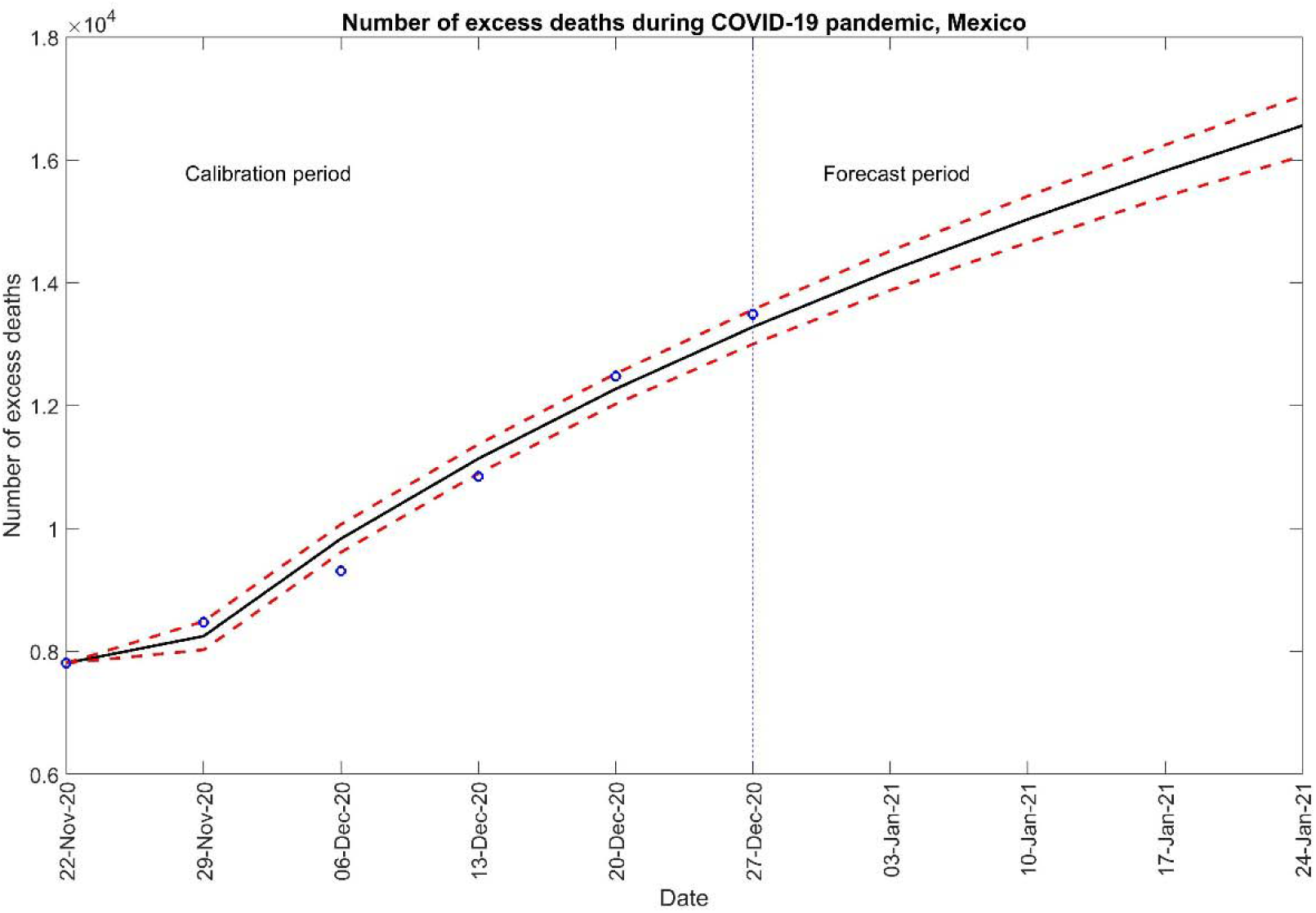
Model-based forecast of excess number of deaths for first 4 weeks of 2021, Mexico. Blue circles are the estimates of excess mortality rate and model fit based on generalized logistic growth model are shown by the black line. The red dashed lines represent the upper and lower bound of 95% prediction interval. The vertical dashed black line denotes the end of calibration period and start of forecasting period.

## Discussion

Monitoring the excess mortality rate during the course of a pandemic is one of the key approaches for evaluating pandemic mortality impact [19]. In this study we characterized the excess mortality impact during COVID-19 pandemic in Mexico from March 1, 2020 to January 2, 2021. The pandemic was associated with an excess mortality rate of 26.10 per 10,000 population (a total of 333,538 excess deaths). Further, COVID-19 laboratory-confirmed deaths comprised only 38.64% of total excess deaths during the studied period. Our findings indicate that the COVID-19 pandemic has exerted a particularly devastating mortality burden on the Mexican population. We found that the all-cause excess-death rate among males was twice as high as the excess death rate among women in Mexico. This finding is in line with the previous studies, indicating that more men die from COVID-19 than women [20-22]. Several factors such as differences in the prevalence of comorbidities [23] as well as risk behaviors such as smoking and drinking [24], frequency of hand washing [25-27] and delays in health care seeking [22] could be contributing to a higher risk of COVID-19 death among males.

We found that both the all-cause excess death rate and the rate ratio were the highest in Mexico City, compared to rest of the country. A previous study reported that Mexico City was the most affected area during 2009-10 A/H1N1 influenza pandemic in Mexico [28]. Mexico City is one of the most crowded cities in the world [29] and has been significantly affected by air pollution for decades [30]. Prior work has identified high population density [31, 32] and long-term exposure to ambient air pollution [33, 34] as significant predictors of COVID-19 death. Besides, the long term exposure to ambient air pollution, and living in overcrowded setting is not random and might interact with other social determinants of health [34] such as poverty, unemployment, and lack of healthcare access that increase the risk of death during natural disasters.

The fraction of COVID-19 attributed excess deaths was lower in Mexico (38.64%) compared to more than 65% in the USA [19, 35], and Germany [36]. From March to May 2020, the number of all cause excess deaths in the US was only 28% higher than the official record of COVID-19 deaths for that period [19]. Subsequently, from March 15, 2020 to January 30, 2021 an estimated 527,500 excess deaths occurred in the USA of which 83.3% were attributed to COVID-19 [37]. In developed countries like Germany where the COVID-19 pandemic management has been considered a success story, the estimated excess number of deaths during the first wave of pandemic was lower than the reported number of COVID-19 deaths (+8071 estimated excess deaths vs. 8674 reported COVID-19 deaths) [36]. The low proportion of COVID-19 deaths out of total excess deaths could be driven by low COVID-19 testing rates in the country [4], delays in reporting COVID-19 deaths [38], diagnostic delays for fatal conditions such as cancer [39], issues related to the sensitivity of reverse transcriptase-PCR (RTPCR) test [40], disruption of routine health care owing to the collapse of the health system, or intentional choices of not visiting health facilities due to fear of contracting the virus, exacerbating the effects on the health of vulnerable groups [41]. Moreover, the pandemic triggered a mental health crisis that has given rise to an increase in self-harm and suicide [42, 43]. Our forecast of excess deaths during the first four weeks of 2021 showed an increasing trend which was in line with the high morbidity trend of COVID-19 cases surrounding that time period [44].

Our Twitter signal indicate an increasing trend in the Twitter chatter about deaths from mid-January 2020 that peaked in mid-February 2020. This increase coincided with a series of events including the declaration of the novel coronavirus outbreak in Wuhan as a public health emergency of international concern (PHEC) (January 30, 2020), and the WHO-China joint mission of experts from different countries to inform planning on next steps in the response to COVID-19 outbreak (16-24 February, 2020) [45]. Following a short period of decline, the Twitter chatter showed a substantial increase, just as the trend in lab-confirmed COVID-19 deaths started to rise in Mexico. However, it substantially declined during the next few months probably due to pandemic fatigue. This normalization of the pandemic also reflects people’s beliefs of the government signaling that the pandemic was subsiding during the second half of 2020 [46, 47].

Several factors could explain the large number of all-cause excess deaths in Mexico (333,538). First, Mexico has a high burden of non-communicable diseases. In 2019, the top 5 leading causes of deaths were ischemic heart disease, diabetes, chronic kidney disease, cirrhosis, and stroke [48]. These comorbidities have been found to be associated with severe outcomes including death due to COVID-19 [49, 50]. Therefore, COVID-19 pandemic in a country like Mexico with high prevalence of chronic diseases, as well as with a health system struggling with absenteeism and health worker infections might have led to this alarming number of excess deaths [51, 52]. It is worth noting that, Mexico has the highest number of health worker deaths due to the COVID-19 pandemic (∼1400 deaths) in the world [53-55].

The COVID-19 pandemic has a higher all cause excess mortality compared to 2009 A/H1N1 pandemic in Mexico [56] as shown in Table 2. Past work has reported estimates of respiratory excess mortality in Mexico City during the 1918-19 influenza pandemic [57]. Our all-cause excess mortality rate of 63.54 for Mexico City was less than the estimated respiratory excess mortality of 72.90 per 10,000 population for the three waves of 1918 influenza pandemic in Mexico City [57] (table 2).

**Table 2.**
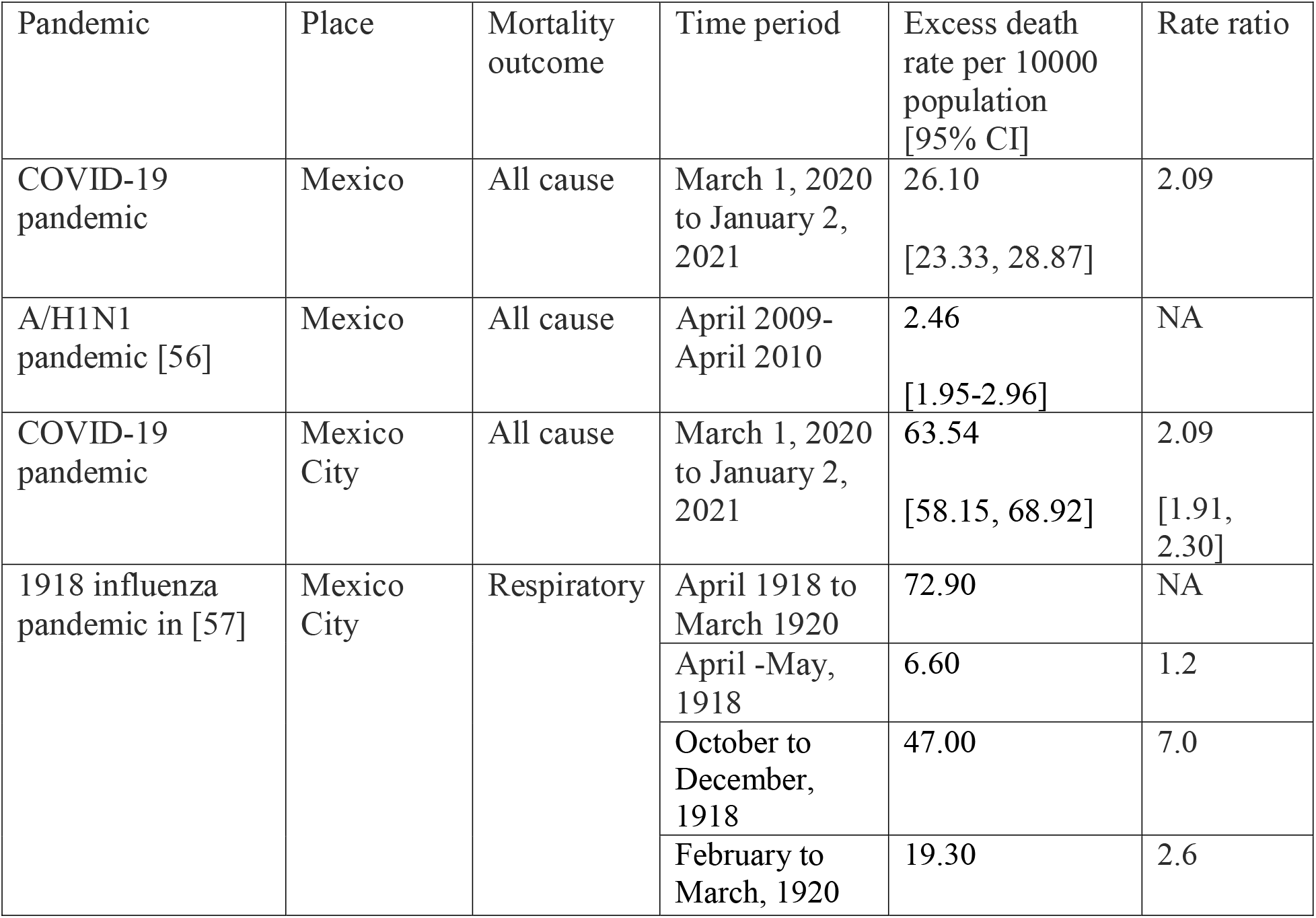
Comparison of excess death rate and rate ratio across different pandemics in Mexico and Mexico City

| Pandemic | Place | Mortality outcome | Time period | Excess death rate per 10000 population [95% CI] | Rate ratio |
| --- | --- | --- | --- | --- | --- |
| COVID-19 pandemic | Mexico | All cause | March 1, 2020 to January 2, 2021 | 26.10<br>[23.33, 28.87] | 2.09 |
| A/H1N1 pandemic [56] | Mexico | All cause | April 2009-April 2010 | 2.46<br>[1.95-2.96] | NA |
| COVID-19 pandemic | Mexico City | All cause | March 1, 2020 to January 2, 2021 | 63.54<br>[58.15, 68.92] | 2.09<br>[1.91, 2.30] |
| 1918 influenza pandemic in [57] | Mexico City | Respiratory | April 1918 to March 1920 | 72.90 | NA |
|  |  |  | April -May, 1918 | 6.60 | 1.2 |
|  |  |  | October to December, 1918 | 47.00 | 7.0 |
|  |  |  | February to March, 1920 | 19.30 | 2.6 |

Our study has several limitations. As excess death rates will be strongly different among subgroups (it is quite high among the elderly, and those with underlying diseases), overall estimate is affected by structure of the population. A detailed data on death certificate with age and underlying diseases information will provide more accurate estimates. Likewise, we can not rule out the possibility of negative excess deaths following this elevated period of excess mortality due to new innovations on vaccination and treatment which might prevent serious complications and deaths, or due to the reduction of the vulnerable populations such as the elderly during the initial pandemic years. Similarly, the COVID-19 deaths data that we have used might be underestimated because of different factors such as very low testing rates in Mexico, and misclassification of COVID-19 deaths. Further studies are needed to shed light on the extent of deaths directly attributable to COVID-19 and those that are related to other causes.

## Conclusion

Our estimate of all-cause excess mortality rate at 26.10 per 10,000 population during COVID-19 pandemic in Mexico provides a reliable estimate of the mortality impact of COVID-19 in a hard-hit Latin American country with a low testing rate. As more refined mortality data becomes available on different sub-groups of the population, further studies on excess mortality could elucidate the mortality impact of the COVID-19 pandemic in Mexico. Our findings indicate that Mexico has been disproportionately affected by the COVID-19 pandemic.

## Data Availability

Data for the study is publicly available.

## Declarations

### Ethics approval and consent to participate

Not Applicable

### Consent for publication

Not Applicable

### Availability of data and material

The datasets used and/or analysed during the current study are available from the corresponding author on reasonable request.

### Author Contributions

Conceptualization, G.C., and S.D.; methodology, G.C., S.D., J.M.B., K.M.; validation, G.C.; formal analysis, S.D.; investigation, S.D.; resources, G.C., S.D., J.M.B.; writing—original draft preparation, S.D.; writing, review and editing, S.D., G.C., J.M.B., A.I.B, K.M. All authors have read and approved the final version of the manuscript.

## Acknowledgements

Not Applicable

